# Effectiveness of new treatment modalities for localized prostate cancer through patient-reported outcome measures: 5 years comparative study

**DOI:** 10.64898/2026.03.04.26347624

**Authors:** Amanda Miranda-Martins, Olatz Garin, José Francisco Suárez, Cristina Gutiérrez, Ferran Guedea, Patricia Cabrera, Manuel Castells, Ismael Herruzo, Lluis Fumadó, Pilar Samper, Carlos Ferrer, Lucas Regis, Ángels Pont, Montse Ferrer, The Multicentric Spanish Group of Clinically Localized Prostate Cancer

**Affiliations:** Health Services Research Group, Hospital del Mar Research Institute, Barcelona, Spain; Universitat Pompeu Fabra (UPF), Barcelona, Spain; CIBER en Epidemiología y Salud Pública, CIBERESP, Spain; Urology Department, Hospital Universitari de Bellvitge, L’Hospitalet de Llobregat, Barcelona, Spain; Urology Department, Hospital del Mar, Barcelona, Spain; Radiation Oncology Department, Institut Català d’Oncologia, L’Hospitalet de Llobregat, Barcelona, Spain; Radiation Oncology Department, Hospital Universitario Virgen del Rocío, Sevilla, Spain; Radiation Oncology Department, Hospital Regional Universitario Carlos Haya, Málaga, Spain; Radiation Oncology Department, Hospital Universitario Rey Juan Carlos, Móstoles, Spain; Radiation Oncology Department, Hospital Provincial de Castellón, Castellón, Spain; Urology Department, Hospital Universitario Vall d’Hebron, Barcelona, Spain

**Keywords:** Localized prostate cancer, Comparative effectiveness, Patient-reported outcome measures, Robot-assisted radical prostatectomy, Intensity-modulated radiotherapy

## Abstract

**Background:** No randomized clinical trial comparing the most established new modalities of treatment for patients with localized prostate cancer has been published, and there is scarce comparative effectiveness research assessing Patient-Reported Outcome Measures (PROMs).

**Objective:** to compare the impact of active surveillance, robot-assisted radical prostatectomy (RARP), Intensity-modulated radiotherapy (IMRT), and real-time brachytherapy on patients, through PROMs, from pre-treatment to five years after diagnosis of localized prostate cancer.

**Methods:** Prospective observational study (ClinicalTrials.gov, NCT05523856) of 566 male patients diagnosed in 2014 to 2021 with clinically localized prostate cancer (50-75 years old; stage cT1 or cT2, N0/Nx and M0/Mx; Gleason ≤ 6 or 7 (if 3 + 4 with T1c); and PSA ≤ 10 ng/ml) and followed until 2019-2026. The Expanded Prostate Cancer Index Composite (EPIC-26) measures urinary incontinence, urinary irritative/obstructive symptoms, sexual, bowel and hormonal domains. EPIC-26 was centrally administered via telephone interviews before treatment and then annually after treatment. Generalized estimating equation (GEE) models were constructed with overlap propensity score-based weights and adjusted by age and clinical tumor stage.

**Results:** Weighted results of adjusted GEE models showed significant declines for sexual health during the 5yr in all treatment groups (ranging from -19.8 to -27.6), but this worsening appeared earlier in those of active treatment (RARP, IMRT and brachytherapy) than in active surveillance. The RARP group presented the greatest deterioration in urinary incontinence (-28.5 vs -11.7 in active surveillance), while the greatest impairment in bowel symptoms was observed in both radiotherapy groups (around -3 vs +0.3 in active surveillance).

**Conclusion:** Our findings provide detailed novel evidence, measured over 5 yr, on the long-term impact of disease and treatment on patients with localized prostate cancer. While all treatment groups showed large sexual deterioration overtime, important differences in urinary incontinence (highest after RARP) and bowel symptoms (after IMRT and brachytherapy) persisted. These findings can inform patients during shared decision-making on the alignment between localized prostate cancer treatment choices and their priorities.

## INTRODUCTION

Prostate cancer is the second most prevalent and the fourth most incident neoplasm among men worldwide (1). Around 75% of patients are diagnosed of cancer localized to the prostate, with an almost 5-year survival rate of 100% (2). The ProtecT (Prostate Testing for Cancer and Treatment) randomized clinical trial showed similar very high survival rates at ten years of follow-up (3) for radical prostatectomy, external radiotherapy and active monitoring, though differing in side effects evaluated with Patient-Reported Outcome Measures (PROMs) (4).

ProtecT patients were treated over 25 years ago (4) with open retropubic radical prostatectomy, external beam 3D-conformal radiotherapy, and initial protocols ofactive monitoring, while nowadays new modalities of them, such as robot-assisted radical prostatectomy (RARP) and intensity-modulated radiation therapy (IMRT), are the mostly being applied.

Metastasis free survival of men in low- or intermediate-risk prostate cancer on active surveillance in 25 cohorts of 15 countries diagnosed from 2000, has increased in the 2013–2016 period, indicating overall safety improvement (5). Studies on RARP presented inconsistent results among techniques, with Retzius-sparing showing favorable early continence outcomes (6,7). IMRT has allowed treating patients with higher doses of radiation while reducing the risk of late gastrointestinal toxicity, enabling biochemical control (8). Real-time brachytherapy enables the precise implementation of radioactive seeds with a steep dose gradient, sparing organs at risk from unnecessary irradiation and minimising treatment time (9).

To the best of our knowledge, no randomized clinical trial comparing some of these most established new modalities of treatment (active surveillance, RARP, IMRT, and real-time brachytherapy) for patients with localized prostate cancer has been published, and there is scarce comparative effectiveness research assessing PROMs (10–16). Four studies compared some of these treatments with follow-ups of 1 (12,16) or 2 years (10,11), but combining modalities of surgery (RARP ranging 87-95%) (10–12) or external radiotherapy (12,16). As far as we know, only the Comparative Effectiveness Analysis of Surgery and Radiation for Localized Prostate Cancer (CEASAR) project provides results of patients treated with the new modalities of the most established treatments followed over 5 years (14,17,18).

Therefore, the aim of this study was to compare the impact of active surveillance, RARP, IMRT, and real-time brachytherapy on patients, through Patient Reported Outcome Measures (PROMs), from pre-treatment to five years after diagnosis of localized prostate cancer, considering side effects (incontinence, irritative or obstructive urinary symptoms, sexual dysfunction, bowel symptoms), as well as mental and physical health.

## MATERIALS AND METHODS

### Patients

The guidelines in Strengthening the Reporting of Observational studies in Epidemiology (STROBE) were followed. This was a prospective observational study (ClinicalTrials.gov Identifier: NCT05523856) involving a cohort of 566 male patients diagnosed with clinically localized prostate cancer provident of the “Multicentric Spanish Group of Clinically Localized Prostate Cancer”, who were diagnosed in 2014 to 2021 and followed over a period of 5 years until 2019-2026. The study details have been described previously (19). Inclusion criteria were 50-75 years old; clinical stage T1 or T2, N0/Nx and M0/Mx; Gleason ≤ 6 or 7 (if 3 + 4 with T1c); Prostate-Specific Antigen (PSA) ≤ 10 ng/ml; and to be treated with active surveillance, RARP, IMRT or real-time brachytherapy as monotherapy.

Decision regarding the initial primary treatment was made jointly by the patients and physician after diagnosis and treatment group assignment was based on the initial primary treatment. This study was approved by the ethics review boards of the 18 participating hospitals (Research Ethics Committee with medicines (CREm) at Bellvitge University Hospital: PR086/ 14), and written informed consent was requested from patients in this study.

### Treatments

Patients undergoing active surveillance were monitored with regular tests including PSA and digital rectal examination on a semiannual basis, and a magnetic resonance imaging and prostate biopsy during the first year. Subsequently, physicians scheduled periodic evaluations based on clinical information, progression and/or physical examination at 6–12 monthly intervals, and repeat biopsy regularly at 1–4 yearly intervals. Transition to alternative treatment modalities were made when Gleason or local TNM progression, significant increase of PSA values and/or PSA doubling time in less than 36 months, or the patient’s choice.

The RARP applied consisted in a prostate extraction procedure carried out through a six-port transperitoneal approach, using the four-arm Da Vinci Si Robotic Surgical System. Nerve-sparing procedure was applied in 90% of the patients, and modifications of the technique including lymphadenectomy were performed only in nine patients according to the final pathological report.

IMRT was carried out with volumetric modulated arc therapy (VMAT) under daily image-guided radiation therapy (IGRT). The planning target volume of the prostate was defined as the entire prostate plus a 5 mm margin in all directions except posteriorly, where a 3 mm margin was used. The treatment was delivered in 3 Gy daily fractions, 5 days per week, with a prescription dose of 60 Gy.

All patients in the real-time interstitial radiotherapy group underwent low-dose-rate brachytherapy with I125 permanent seeds implantation, with a prescribed dose of 145 Gy to the target volume. The prescription dose (V100) applied in the dosimetry prostate volume was at least 95 %, and the dose received by 90% of the prostate (D90) was 100 %.

### Patient-Reported Outcome Measures

To evaluate the impact of treatment side effects, the Expanded Prostate Cancer Index Composite (EPIC-26) (20,21) and Short Form-36 Healthy Survey (SF-36) (22– 24) were administered before treatment and annually for five years. To ensure homogeneous evaluation, telephone interviews were centrally performed by a trained interviewer.

EPIC-26 evaluates urinary, bowel, sexual, and hormonal domains, with 26 items and scores ranging from 0 to 100 (better outcomes) (25). Differences were considered to be of clinical significance if they exceed the following minimum clinically important differences (MCIDs) for the EPIC scores (26): 6-9 points in urinary incontinence; 5-7 in urinary irritative-obstructive symptoms; 10-12 in sexual; 4-6 in bowel; and 4-6 in hormonal summaries. In addition to EPIC scores, one key EPIC item per domain previously selected in the ProtecT study (4) was dichotomized to reflect the proportion of men reporting any problem.

SF-36 generates physical and mental component summaries (PCS and MCS) standardized to have a mean of 50 and standard deviation (SD) of ten In US population (22,23).

The sample size calculated to detect small differences between groups (0.3 SD) on the EPIC or SF-36v2 scores was of 90 patients per treatment group, given a statistical power of at least 80% at a significance level of 5%, and loss to follow-up of 10%.

### Statistical analysis

To account for treatment selection bias, overlap propensity score weighting was performed. The R package WeightIt was used to estimate propensity scores from generalized boosted models (GBM), and to apply the overlap propensity score weighting method, by weighting to target the average treatment effect in the overlap population (ATO). Covariate balance after weighting was evaluated using standardized mean differences (27). Multiple imputation was used to handle missing data for variables included in the model.

Summary statistics for patient characteristics at diagnosis were estimated as unweighted and weighted, with weights applied using complex survey methods. To show unweighted results of EPIC-26 and SF-36 scores boxplots were constructed. Figures were constructed to show the unweighted percentage of men reporting problems in the selected EPIC items, their 95% confidence intervals (95%Cis) calculated using the exact Clopper-Pearson binomial method, cross-sectional differences evaluated with χ^2^ tests, and temporal changes (treatment-by-time interaction) assessed using Wald χ^2^ tests.

To assess PROM changes over time, while accounting for correlation among repeated measures, separate generalized estimating equation (GEE) weighted models were constructed for each EPIC-26 and SF-36 score as dependent variables. Treatment (with active surveillance as a reference group), time, and their interaction were included in the models. The bootstrapping method was used to assess uncertainty in the sampling distribution for the EPIC-26 and SF-36 mean scores. All analysis were performed using R v4.2.2 (R Foundation for Statistical Analysis, Vienna, Austria).

## Results

Of the 566 participants, 87 were treated with active surveillance, 194 with RARP, 107 with IMRT and 178 with pre planned low-dose-rate brachytherapy. At 5 years after treatment, 20 had died, 25 were lost of follow up, and 6 missed the interview. Of the 546 patients surviving, 515 completed PROM data at 5 years (94% completion rate). Median follow-up was 60.5 months (interquartile range 60.0-62.6). Of 87 patients initially under active surveillance, 17 initiated treatment with: RARP (4), IMRT (11), brachytherapy (1), and IMRT combined with brachytherapy (1).

**Table 1** shows unweighted and weighted estimates of clinical characteristics. Although most of these unweighted estimates presented statistically significant differences among treatment groups, after applying overlap propensity score-based weights, all were well balanced except for: clinical tumor stage (T1c 71.5%-100%, *p*<0.001) and age (mean 62.8-66.8, *p*=0.007).

**Table 1.**
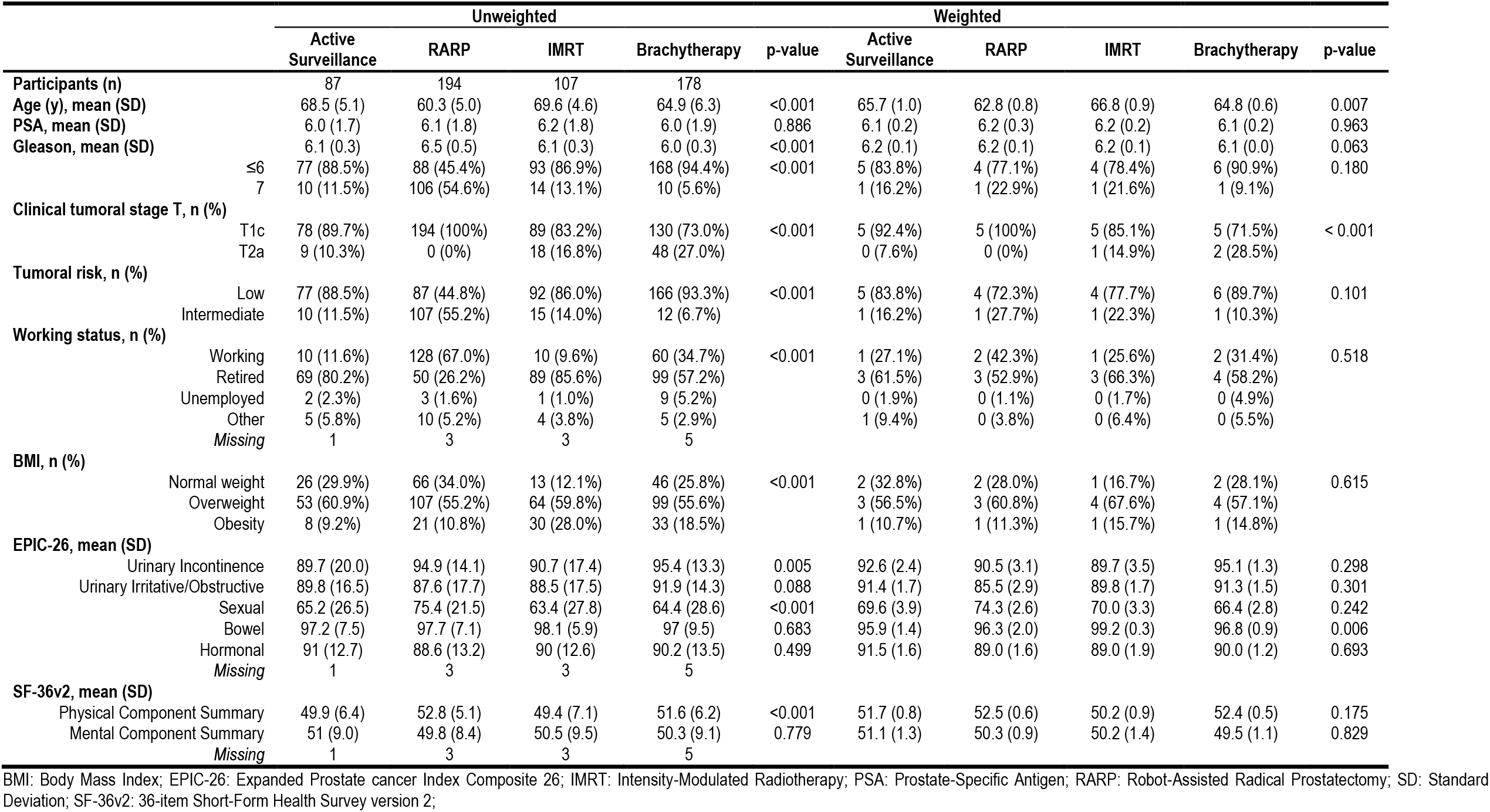
Unweighted and weighted descriptive of patient characteristics and quality of life scores before treatment (n=566).

Figure 1. shows the unweighted results of the EPIC-26 urinary domain. Patients treated with RARP experienced the largest and persistent decline in urinary continence scores through the 5 years follow up **(Figure 1A)**, with median scores remaining around 70 compared with medians of 100 in the other treatment groups. The use of absorbent pad followed a similar pattern **(Figure 1B)**, with approximately 25–35% of patients in the RARP group reporting use of ≥1 pad per day compared with lower than 5% in the other groups (p < 0.001). Urinary irritative/obstructive domain scores remained relatively stable across treatment groups **(Figure 1C)**, and the percentage of patients reporting pain or burning on urination only increased at 1st year among patients treated with brachytherapy **(Figure 1D)**.

**Figure 1.**
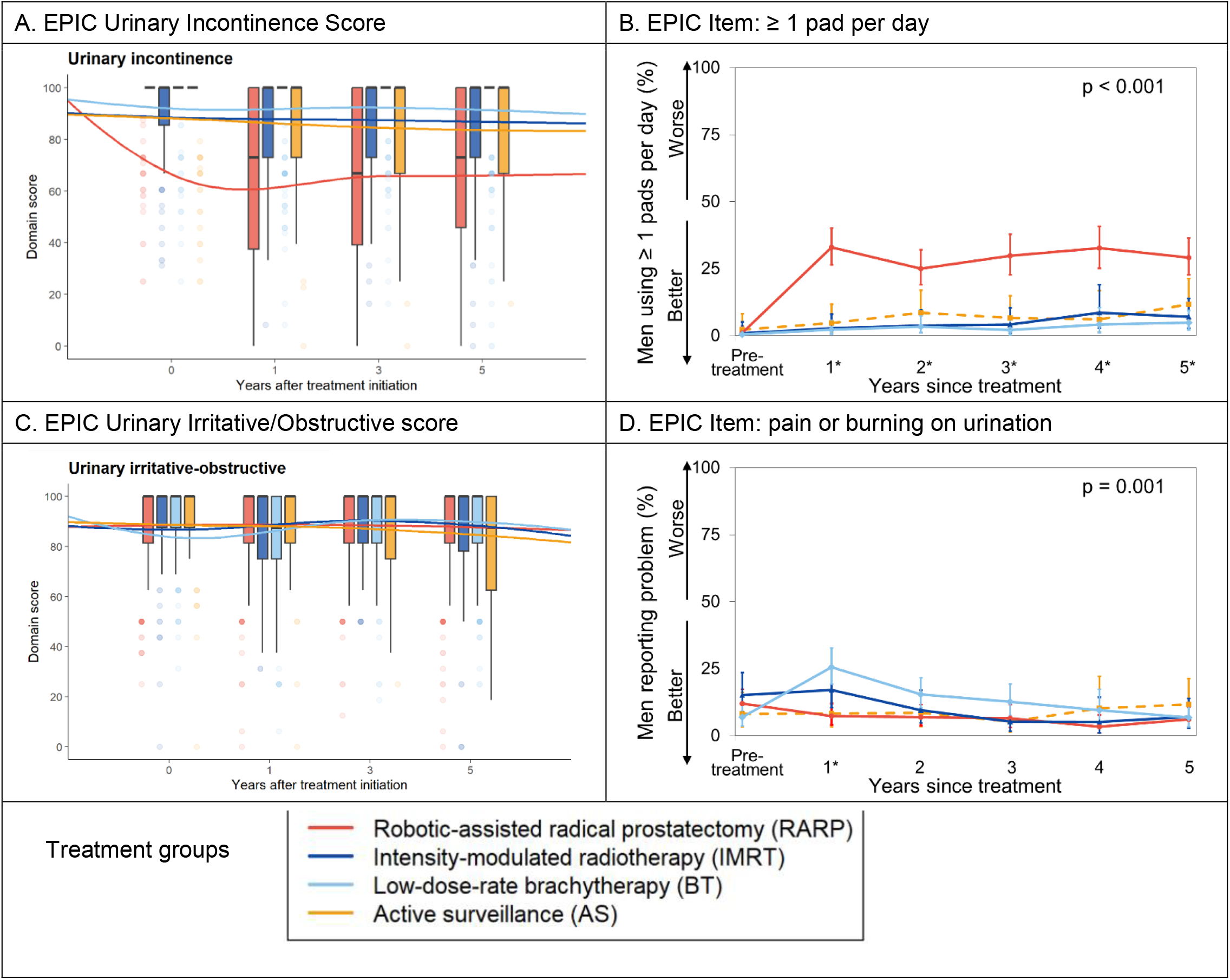
Unweighted five years follow-up results of the EPIC-26 urinary domain. **Footnote:** Boxplots of EPIC-26 urinary scores (range 0–100, higher scores indicate better outcomes) are display in Panels A) urinary incontinence and C) urinary irritative/obstructive domain scores, at pretreatment and at 1, 3, and 5 years after diagnosis. Boxes represent the interquartile range (IQR), lines inside the boxes indicate the median, whiskers extend to the most extreme values within 1.5 × IQR. All the points beyond 1.5 × interquartile ranges are shown as dots, the intensity of which signifies the relative number of participants with that value. Panels B and D present the percentage of problems for selected EPIC-26 items: error bars indicate 95% confidence intervals, and p values correspond to temporal changes (treatment-by-time interaction) assessed using Wald χ^2^ tests.

Sexual scores declined in all treatment groups through follow-up **(Figure 2A**). The percentage of men reporting erection not firm for intercourse increased progressively in all groups during the 5-year follow-up period, remaining lower than 50% in patients treated with RARP and at its highest level of 63-84% in patients treated with IMRT **(Figure 2B**). Bowel and hormonal scores were stable across groups through all evaluations, indicating minimal symptom burden **(Figure 2C and 2E**). Men reporting problems with fecal incontinence or hot flashes were rare (**Figure 2D and F**).

**Figure 2.**
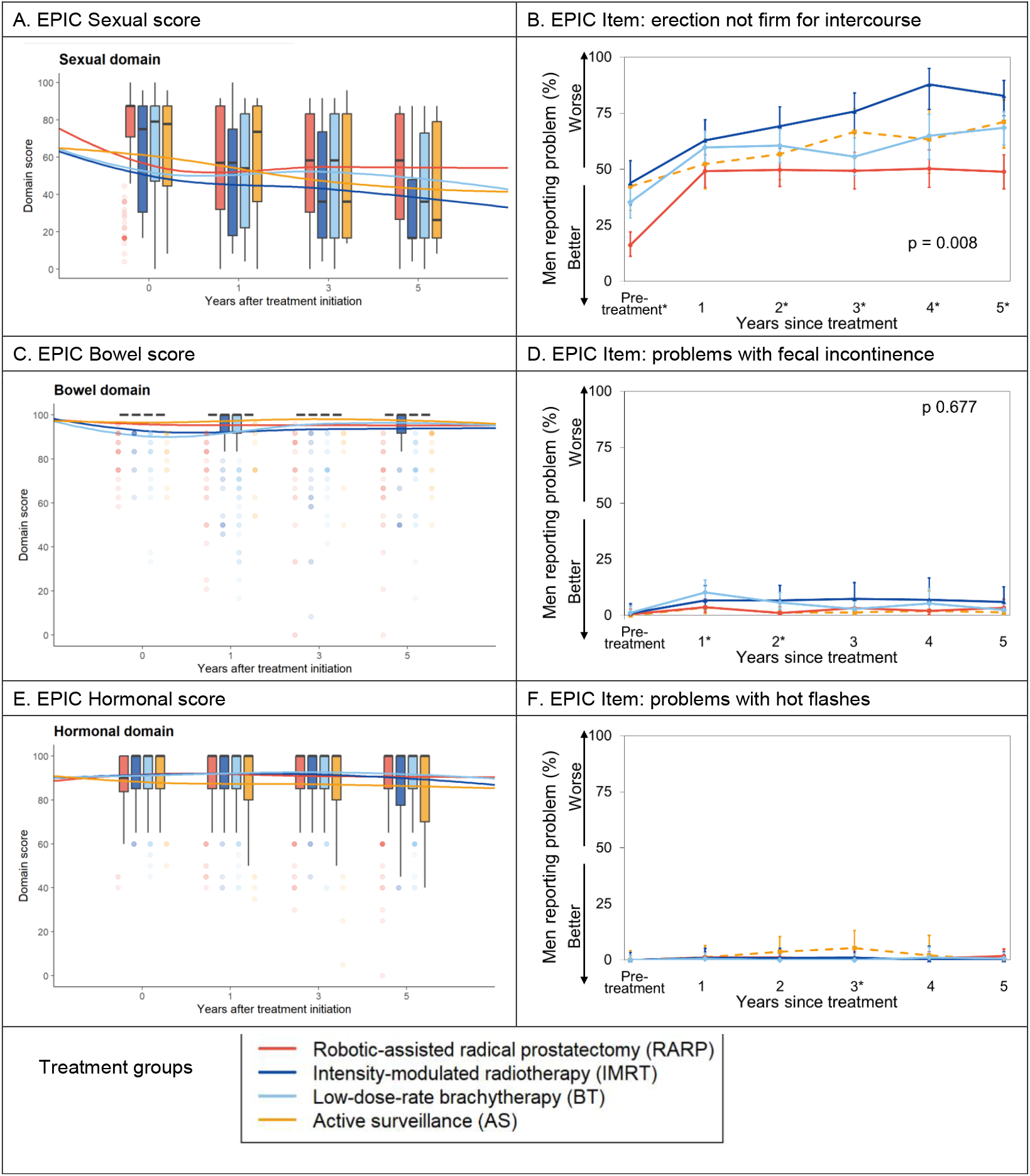
Unweighted five years follow-up results of the EPCI-25 sexual and bowel domains. **Footnote:** Boxplots of EPIC-26 sexual and bowel scores (range, 0–100; higher scores indicate better outcomes) are display in Panels A) Sexual Domain; C) Bowe Domain; and E) Hormonal Domain, at pretreatment and at 1, 3, and 5 years after treatment initiation. Boxes represent the interquartile range (IQR), lines inside the boxes indicate the median, whiskers extend to the most extreme values within 1.5 × IQR. All the points beyond 1.5 × interquartile ranges are shown as dots, the intensity of which signifies the relative number of participants with that value. Panels B, D and F present the percentage of problems for selected EPIC-26 items: error bars indicate 95% confidence intervals, and p values correspond to temporal changes (treatment-by-time interaction) assessed using Wald χ^2^ tests.

**Figure 3A** shows a modest decline over time of the median SF-36v2 physical component summary scores in all groups, whereas mental component summary scores medians remained stable **(Figure 3B)**. For both components, medians’ scores remained around 50, approximating general population reference norms.

**Figure 3.**
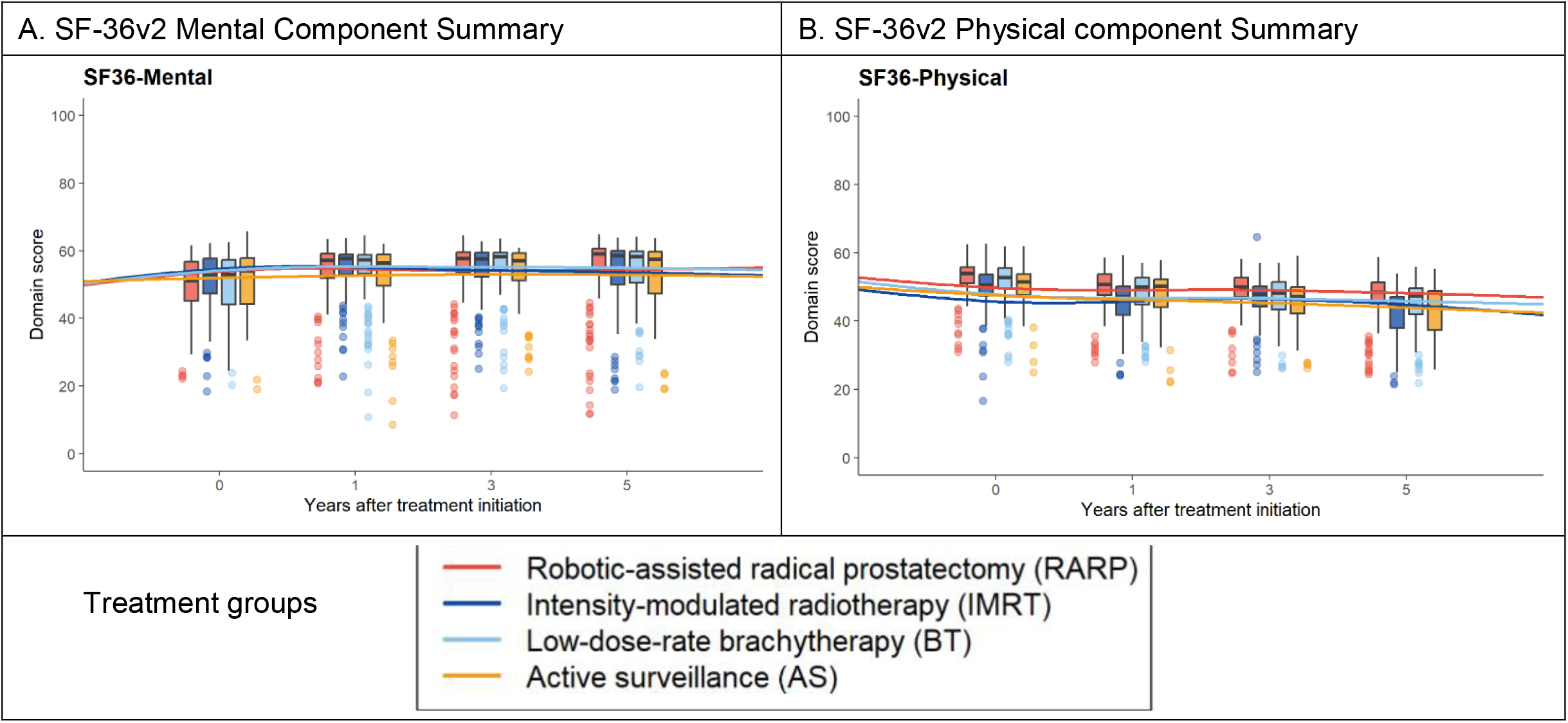
Unweighted five years follow-up results of the SF-36v2 physical and mental health component. **Footnote:** Boxplots of SF-36v2 of mental and physical scores (range, 0–100; higher scores indicate better outcomes) are display in Panels A) Physical Component Summary; and B) Mental Component Summary, at pretreatment and at 1, 3, and 5 years after treatment initiation. Boxes represent the interquartile range (IQR), lines inside the boxes indicate the median, whiskers extend to the most extreme values within 1.5 × IQR. All the points beyond 1.5 × interquartile ranges are shown as dots, the intensity of which signifies the relative number of participants with that value.

**Table 2** shows the mean scores of the EPIC and SF-36v2 estimated through GEE models constructed applying overlap propensity score–weights and adjusted by clinical tumor stage and age. Figure 4 visually summarizes these EPIC mean scores across treatment groups.

**Table 2.** Results of EPIC-26 over time across different treatment groups. estimated with GEE models: means weighted applying propensity scores and adjusted by clinical tumour stage and age.

|  | Urinary Incontinence |  | Irritative/Obstructive |  | Bowel |  | Sexual |  | Hormonal |  | SF-36v2 PCS |  | SF-36v2 MCS |  |
| --- | --- | --- | --- | --- | --- | --- | --- | --- | --- | --- | --- | --- | --- | --- |
|  | Adjusted mean | 95% CI | Adjusted mean | 95% CI | Adjusted mean | 95% CI | Adjusted mean | 95% CI | Adjusted mean | 95% CI | Adjusted mean | 95% CI | Adjusted mean | 95% CI |
| **Active Surveillance** |  |  |  |  |  |  |  |  |  |  |  |  |  |  |
| Pre-treatment | 92.8 | [ 89.3 · 96.4 ] | 90.5 | [ 87.5 · 93.5 ] | 96.2 | [ 94.1 · 98.7 ] | 71.0 | [ 64.8 · 78.0 ] | 91.7 | [ 88.9 · 94.8 ] | 52.0 | [ 50.6 · 53.4 ] | 50.9 | [ 48.8 · 52.8 ] |
| 1 year | 91.1 | [ 87.5 · 94.8 ] | 90.3 | [ 87.3 · 93.3 ] | 96.9 | [ 95.1 · 98.9 ] | 70.7 | [ 64.2 · 76.1 ] | 91.5 | [ 87.4 · 95.4 ] | 50.2 <sup>a</sup> | [ 48.8 · 51.5 ] | 53.2 <sup>a</sup> | [ 50.7 · 55.7 ] |
| 2 year | 91.8 | [ 88.1 · 95.6 ] | 93.0 | [ 90.0 · 96.2 ] | 99.6 <sup>a</sup> | [ 98.8 · 100.4 ] | 66.6 | [ 60.6 · 72.3 ] | 94.2 | [ 91.4 · 97.0 ] | 49.0 <sup>a</sup> | [ 48.0 · 50.2 ] | 55.8 <sup>a</sup> | [ 54.3 · 57.2 ] |
| 3 year | 89.2 | [ 83.4 · 95.2 ] | 88.3 | [ 83.8 · 93.1 ] | 98.8 <sup>a</sup> | [ 98.1 · 99.6 ] | 59.1 <sup>a</sup> | [ 51.6 · 66.3 ] | 92.8 | [ 89.3 · 96.5 ] | 47.5 <sup>a</sup> | [ 46.5 · 48.7 ] | 55.2 <sup>a</sup> | [ 53.1 · 57.3 ] |
| 4 year | 88.0 | [ 80.4 · 95.0 ] | 88.1 | [ 84.0 · 92.1 ] | 98.2 | [ 96.9 · 99.6 ] | 59.4 <sup>a</sup> | [ 50.7 · 67.6 ] | 92.4 | [ 89.0 · 95.6 ] | 48.4 <sup>a</sup> | [ 46.9 · 49.7 ] | 53.5 <sup>a</sup> | [ 51.3 · 55.6 ] |
| 5 year | 81.1 <sup>a</sup> | [ 75.2 · 88.3 ] | 78.9 <sup>a</sup> | [ 73.8 · 84.9 ] | 96.5 | [ 94.9 · 98.5 ] | 44.9 <sup>a</sup> | [ 36.3 · 53.0 ] | 86.0 <sup>a</sup> | [ 81.4 · 90.2 ] | 44.2 <sup>a</sup> | [ 42.5 · 45.9 ] | 51.5 | [ 48.6 · 54.1 ] |
| **RARP** |  |  |  |  |  |  |  |  |  |  |  |  |  |  |
| Pre-treatment | 90.2 | [ 86.0 · 95.4 ] | 83.3 <sup>b</sup> | [ 79.0 · 88.3 ] | 96.1 | [ 93.6 · 99.2 ] | 71.1 | [ 66.4 · 75.3 ] | 88.2 | [ 85.8 · 90.9 ] | 52.1 | [ 50.9 · 53.1 ] | 50.4 | [ 48.6 · 52.3 ] |
| 1 year | 67.5 <sup>a,b,c</sup> | [ 61.0 · 74.2 ] | 84.8 | [ 76.6 · 93.5 ] | 96.4 | [ 94.4 · 98.7 ] | 48.7 <sup>a,b,c</sup> | [ 41.6 · 55.2 ] | 89.4 | [ 84.4 · 94.8 ] | 48.1 <sup>a</sup> | [ 46.4 · 49.8 ] | 53.1 | [ 49.8 · 56.6 ] |
| 2 year | 69.4 <sup>a,b,c</sup> | [ 63.5 · 75.8 ] | 88.8 | [ 82.1 · 96.1 ] | 93.2 <sup>b,c</sup> | [ 89.2 · 98.3 ] | 51.0 <sup>a,b,c</sup> | [ 44.6 · 56.9 ] | 87.1 <sup>b</sup> | [ 82.5 · 92.6 ] | 46.5 <sup>a</sup> | [ 44.0 · 49.3 ] | 52.7 | [ 49.9 · 55.9 ] |
| 3 year | 64.1 <sup>a,b,c</sup> | [ 55.2 · 73.6 ] | 82.6 | [ 76.6 · 90.1 ] | 94.5 | [ 90.3 · 99.1 ] | 49.5 <sup>a</sup> | [ 42.5 · 56.3 ] | 86.4 <sup>b</sup> | [ 82.3 · 91.4 ] | 46.0 <sup>a</sup> | [ 43.2 · 49.6 ] | 50.9 <sup>b</sup> | [ 48.8 · 53.6 ] |
| 4 year | 69.5 <sup>a,b</sup> | [ 58.7 · 80.0 ] | 86.0 | [ 82.3 · 90.6 ] | 97.8 | [ 96.2 · 99.3 ] | 45.9 <sup>a,b</sup> | [ 38.3 · 52.9 ] | 89.2 | [ 85.0 · 94.6 ] | 45.4 <sup>a</sup> | [ 42.7 · 48.9 ] | 53.0 | [ 50.5 · 56.2 ] |
| 5 year | 61.7 <sup>a,b,c</sup> | [ 54.4 · 70.6 ] | 77.7 | [ 70.8 · 86.8 ] | 94.2 | [ 90.6 · 98.8 ] | 43.5 <sup>a</sup> | [ 36.9 · 50.0 ] | 87.2 | [ 82.3 · 93.4 ] | 45.2 <sup>a</sup> | [ 43.4 · 47.3 ] | 51.1 | [ 47.4 · 56.4 ] |
| **IMRT** |  |  |  |  |  |  |  |  |  |  |  |  |  |  |
| Pre-treatment | 88.1 | [ 82.6 · 94.1 ] | 88.4 | [ 85.0 · 91.8 ] | 99.7 <sup>b</sup> | [ 98.4 · 100.9 ] | 69.7 | [ 65.0 · 74.3 ] | 89.4 | [ 86.1 · 92.5 ] | 50.7 | [ 49.2 · 52.2 ] | 49.7 | [ 47.5 · 52.0 ] |
| 1 year | 85.2 | [ 77.9 · 94.4 ] | 88.1 | [ 84.0 · 92.7 ] | 92.3 <sup>a,b,c</sup> | [ 89.3 · 95.2 ] | 56.0 <sup>a,b</sup> | [ 48.6 · 63.3 ] | 92.8 | [ 89.9 · 95.9 ] | 47.8 <sup>a,b</sup> | [ 46.4 · 49.1 ] | 54.6 <sup>a</sup> | [ 53.2 · 56.1 ] |
| 2 year | 85.9 | [ 79.1 · 94.1 ] | 87.4 | [ 82.3 · 93.5 ] | 95.0 <sup>a,b,c</sup> | [ 92.6 · 97.5 ] | 52.3 <sup>a,b,c</sup> | [ 46.1 · 58.4 ] | 91.2 | [ 88.1 · 94.0 ] | 47.1 <sup>a,b</sup> | [ 45.7 · 48.5 ] | 55.5 <sup>a</sup> | [ 54.1 · 57.0 ] |
| 3 year | 83.9 | [ 76.8 · 92.1 ] | 87.7 | [ 83.7 · 92.5 ] | 92.5 <sup>a,b,c</sup> | [ 88.8 · 96.3 ] | 48.5 <sup>a</sup> | [ 40.4 · 55.4 ] | 93.1 | [ 90.1 · 96.1 ] | 47.7 <sup>a</sup> | [ 46.4 · 48.9 ] | 54.6 <sup>a</sup> | [ 52.7 · 56.5 ] |
| 4 year | 86.4 | [ 79.7 · 94.2 ] | 90.2 | [ 86.7 · 94.0 ] | 93.0 <sup>a,c</sup> | [ 88.8 · 98.2 ] | 38.9 <sup>a,b,c</sup> | [ 29.8 · 47.3 ] | 89.1 | [ 83.6 · 94.6 ] | 45.7 <sup>a,b</sup> | [ 44.2 · 47.3 ] | 55.8 <sup>a</sup> | [ 54.0 · 57.5 ] |
| 5 year | 83.5 | [ 77.2 · 91.2 ] | 87.4 <sup>b,c</sup> | [ 83.8 · 91.5 ] | 96.5 <sup>a,c</sup> | [ 94.6 · 98.2 ] | 42.8 <sup>a</sup> | [ 35.5 · 49.7 ] | 92.3 <sup>b</sup> | [ 89.2 · 95.3 ] | 44.9 <sup>a</sup> | [ 43.5 · 46.2 ] | 55.4 <sup>a,b,c</sup> | [ 53.8 · 57.0 ] |
| **Brachytherapy** |  |  |  |  |  |  |  |  |  |  |  |  |  |  |
| Pre-treatment | 95.2 | [ 93.3 · 97.3 ] | 92.6 | [ 90.1 · 95.3 ] | 97.3 | [ 95.4 · 98.9 ] | 65.2 | [ 60.8 · 69.6 ] | 90.4 | [ 88.4 · 92.5 ] | 52.5 | [ 51.7 · 53.3 ] | 49.6 | [ 48.0 · 51.4 ] |
| 1 year | 91.1 <sup>a</sup> | [ 87.9 · 94.3 ] | 83.4 <sup>a,b,c</sup> | [ 79.8 · 87.2 ] | 89.5 <sup>a,b,c</sup> | [ 86.7 · 92.5 ] | 53.5 <sup>a,b</sup> | [ 49.1 · 57.8 ] | 91.7 | [ 89.7 · 94.1 ] | 48.3 <sup>a,b,c</sup> | [ 47.4 · 49.3 ] | 53.9 <sup>a</sup> | [ 52.7 · 55.3 ] |
| 2 year | 93.5 | [ 90.5 · 96.6 ] | 87.1 <sup>a,b,c</sup> | [ 84.0 · 90.5 ] | 93.1 <sup>a,b,c</sup> | [ 90.5 · 95.9 ] | 51.5 <sup>a,b</sup> | [ 47.5 · 55.4 ] | 90.5 | [ 88.1 · 92.8 ] | 47.1 <sup>a,b,c</sup> | [ 46.3 · 48.0 ] | 54.2 <sup>a</sup> | [ 52.9 · 55.5 ] |
| 3 year | 92.4 | [ 89.3 · 95.9 ] | 92.6 | [ 89.6 · 95.8 ] | 94.4 <sup>b,c</sup> | [ 92.2 · 96.8 ] | 50.9 <sup>a</sup> | [ 46.0 · 55.9 ] | 92.8 | [ 89.6 · 96.4 ] | 47.1 <sup>a</sup> | [ 46.2 · 47.9 ] | 55.6 <sup>a</sup> | [ 54.1 · 57.0 ] |
| 4 year | 91.6 <sup>a</sup> | [ 89.0 · 94.3 ] | 89.9 | [ 85.9 · 93.9 ] | 94.5 <sup>b,c</sup> | [ 92.2 · 97.1 ] | 41.3 <sup>a,b</sup> | [ 35.6 · 46.6 ] | 92.3 | [ 89.4 · 95.1 ] | 46.1 <sup>a,b,c</sup> | [ 44.9 · 47.2 ] | 56.3 <sup>a,b</sup> | [ 54.6 · 57.8 ] |
| 5 year | 91.0 <sup>a,b</sup> | [ 87.8 · 94.0 ] | 90.3 <sup>b,c</sup> | [ 86.8 · 93.7 ] | 93.9 <sup>a</sup> | [ 91.7 · 96.2 ] | 45.4 <sup>a</sup> | [ 41.1 · 50.0 ] | 90.2 | [ 87.8 · 93.1 ] | 45.4 <sup>a</sup> | [ 44.3 · 46.5 ] | 54.4 <sup>a</sup> | [ 53.1 · 55.9 ] |

a: p-value < 0.05 in the comparison with values before treatment; b: p-value < 0.05 in the comparison with active surveillance (reference group) at each evaluation; c: p-value < 0.05 in difference of change comparing with active surveillance (reference group). 95% CI: 95% Confidence Interval; IMRT: Intensity-Modulated Radiotherapy; MCS: Mental Component Summary; PCS: Physical Component Summary; RARP: Robot-Assisted Radical Prostatectomy.

Patients in the active surveillance group experienced gradual declines in urinary, sexual and SF-36 PCS scores. After RARP, the deterioration of urinary incontinence was statistically higher than in the active surveillance group throughout the 5 years; the sexual score decline was also statistically higher during early follow-up, although between-group differences attenuated over time.

Both radiotherapy groups showed greater declines in bowel function than active surveillance, and maintained urinary incontinence and hormonal scores quite stable. Compared with active surveillance, the urinary irritative/obstructive score presented statistically greater early decline in the brachytherapy group, while sexual decline was higher in the IMRT group.

All treatment groups presented statistically significant decline over time in the SF-36v2 physical component summary scores, whereas mental component summary scores slightly improved.

## DISCUSSION

This is one of the first 5-yr follow-up studies to provide valuable insights into the long-term impact from patients’ perspective of active surveillance, RARP, IMRT, and real-time brachytherapy on urinary, sexual, bowel, and hormonal side effects and on physical and mental health. Weighted results of adjusted GEE models showed significant declines for sexual and physical health during the 5 years in all treatment groups, but worsening of sexual scores appeared earlier in those of active treatment (RARP, IMRT and brachytherapy) than in active surveillance. The RARP group presented the greatest deterioration in urinary incontinence, while the greatest impairment in bowel symptoms was observed in both radiotherapy groups.

In our study, urinary continence exhibited a persistent and large difference on deterioration between treatments, being RARP the single one demonstrating a sustained disadvantage in comparison with active surveillance at 5 years (-28.5 vs -11.7), which was over the MCID of 6-9 points for this domain (26). This pattern aligns with results 5 years after treatment on men with favorable-risk prostate cancer of the CEASAR study (13), estimating -10.9 (95%CI -14.2 to -7.6) points of change difference between these treatments. Although significantly better urinary continence outcomes after RARP with Retzius-sparing technique were reported (6), no comparison could be done since there were only studies with follow-ups until 1 year after treatment (28,29).

Also consistently with CEASAR study among men with favorable-risk prostate cancer (13) results in urinary irritative/obstructive domain, scores remained high regardless of the treatment applied in our study. Only the decline observed in our study for active surveillance at the 5 years assessment drifted from this pattern: -11.6 points of change from pre-treatment, which was over the MCID of 5-7 points defined for this domain (26). This decline could be partially explained by active treatments applied time after the initiation of active surveillance.

Bowel symptoms presented small declines after IMRT and brachytherapy compared to active surveillance, which was around -3 points at the 5th year, but were just below the 4-6 points established as the MCID for this domain (26). Men with favorable-risk prostate cancer from CEASAR study (13) exhibit similar statistically significant differences of change from active surveillance for external radiotherapy (-2.7 at 5th year) and for brachytherapy (-2.3 at 5th year). In both studies, neither active surveillance nor RARP presented any worsening through the follow-up.

The three active treatments assessed in this study demonstrated comparable trajectories in sexual domain, patients experienced the sharpest early decline, which remained over the MCID (10-12 points) (26) until the evaluation at the 5^th^ year: -27.6 in RARP group, -26.9 in IMRT and -19.8 in brachytherapy. In contrast, patients under active surveillance presented stable sexual results until the fourth year, and the decline between the fourth and fifth year of follow-up was large (-14.5 points). Subsequently, the mean scores of all groups were comparable at the end of our study, ranging from 42.8 to 45.4, and quite consistent with CEASAR study results (13).

Our hormonal results aligns with findings from the CEASAR study among men with favorable-risk prostate cancer (13) showing stability, with differences below the MCID of 4-6 (26).

The SF-36v2 physical health component demonstrated a decline of moderate magnitude (more than 0.5 SD) throughout the 5 year follow-up in all groups of our study. This finding differs from those of men with favorable-risk prostate cancer from CEASER study, where the SF-36 physical functioning domain remained stable during the whole follow-up period. However, it is remarkable the content differences between the physical component summary and the physical functioning domain of the SF-36, which could explain partly this. The SF-36v2 mental health component stability across all groups throughout the 5 years follow-up found in our study was quite comparable with results of studies using CEASER population reporting stable emotional wellbeing (18,30). Evidence from a prostate cancer multi-institutional 1 year post treatment study further suggests that baseline psychosocial and clinical factors, rather than treatment choice, are the main determinants of mental well-being after localized prostate cancer (31).

The main limitation of this study lies in its observational design, which restricts causal interpretation of the estimated treatment effects. As in most comparative-effectiveness analyses of localized prostate cancer, treatment selection bias may persist because of clinical, demographic, and personal preferences influence therapy selection. Active surveillance is typically reserved for men with low-risk tumors or advanced age (88.5% and mean age of 68.5 years), whereas RARP is more often applied to younger healthier patients (mean age of 60.3 years). In our cohort, overlap propensity score weights were used to mitigate this treatment selection bias, and residual confounding was addressed adjusting by age and tumour stage the GEE models.

The strengths of the study include the collection of data per the present recommendations of the International Consortium for Health Outcomes Measurement for localized prostate cancer (32), and regular follow-up with high rates of response (94% completion rate at 5 years), and the consistency of interpretation based on the magnitude of change with previously established minimum clinically important differences (MCIDs) for the EPIC scores (26).

## CONCLUSIONS

This study provides a comprehensive five-year evidence of patient-reported outcomes for contemporary treatments of localized prostate cancer. Our findings support that, real-time low-dose rate brachytherapy and IMRT are the treatment options causing the least impact on incontinence and irritative-obstructive symptoms of the urinary domain. Furthermore, brachytherapy also presented the lowest sexual deterioration. The largest deterioration of urinary incontinence was observed in patients underwent RARP, while these patients and those under active surveillance did not present bowel symptoms impairment. These results provide patients, clinicians, and health care planners with clear information to make evidence-based decisions and facilitate shared clinical decision-making, taking into account patients’ perspective.

## Data Availability

The datasets generated and/or analyzed during the current study are available from the corresponding author on reasonable request.

## Acknowledgments

We acknowledge the collaboration of the other members of the Multicentric Spanish Group of Clinically Localized Prostate Cancer for their contributions in data collection, critical revision of the manuscript, and administrative, technical, and material support: Montse Ferrer, Àngels Pont, Olatz Garin, Yolanda Pardo, Víctor Zamora, Mónica Ávila, Cristina Gutiérrez, Ferran Guedea, Montse Ventura, Ferran Ferrer, Ana Boladeras, Andrea Slocker, Miguel Ángel Berenguera, Joan Pera, José Francisco Suárez, Manuel Castells, Elena López, Patricia Cabrera, Juan Manuel Conde, Belén Congregado, Rafael Medina, Ismael Herruzo, Sabrina López, Víctor Baena, José López Torrecilla, Jorge Pastor, Víctor Muñoz, Manuel Enguix, Àlvar Roselló, Arantxa Eraso, Carlos Ferrer, Ángel Sánchez, Francisco Gómez-Veiga, Víctor Macías, Lluís Fumadó, José María Abascal, Josep Jové, Moisés Mira, M^a^ Elena García, Gemma Sancho, Ana Celma, Lucas Regis, Pilar Samper, Luís A Glaría, M^a^ Ángeles Cabeza, Germán Juan, Amalia Palacios, Amelia Béjar, Sonia García. We also thank the patients and families who made this study possible and the clinical study teams who participated in the study. Moreover, the group thanks Áurea Martín for her support in English editing, proofreading, and preparing this manuscript for submission.

## REFERENCES

1. Bray F, Laversanne M, Sung H, Ferlay J, Siegel RL, Soerjomataram I, et al. Global cancer statistics 2022: GLOBOCAN estimates of incidence and mortality worldwide for 36 cancers in 185 countries. CA Cancer J Clin. 2024;74(3):229–63. doi:10.3322/caac.21834

2. Raychaudhuri R, Lin DW, Montgomery RB. Prostate Cancer: A Review. JAMA. 2025 Apr 22;333(16):1433–46. doi:10.1001/jama.2025.0228

3. Hamdy FC, Donovan JL, Lane JA, Mason M, Metcalfe C, Holding P, et al. 10-Year Outcomes after Monitoring, Surgery, or Radiotherapy for Localized Prostate Cancer. N Engl J Med. 2016 Oct 13;375(15):1415–24. doi:10.1056/NEJMoa1606220

4. Donovan JL, Hamdy FC, Lane JA, Mason M, Metcalfe C, Walsh E, et al. Patient-Reported Outcomes after Monitoring, Surgery, or Radiotherapy for Prostate Cancer. N Engl J Med. 2016 Oct 13;375(15):1425–37. doi:10.1056/NEJMoa1606221

5. Bangma C, Doan P, Zhu L, Remmers S, Nieboer D, Helleman J, et al. Has Active Surveillance for Prostate Cancer Become Safer? Lessons Learned from a Global Clinical Registry. Eur Urol Oncol. 2025 Apr 1;8(2):324–37. doi:10.1016/j.euo.2024.07.003

6. Lv T, Yang J, Cheng B. Oncological and functional outcomes of Retzius-sparing vs. standard robot-assisted radical prostatectomy: evidence on randomized-controlled trials studies. J Robot Surg. 2025 Apr 21;19(1):165. doi:10.1007/s11701-025-02335-z PubMed PMID: 40257521.

7. Checcucci E, Veccia A, Fiori C, Amparore D, Manfredi M, Di Dio M, et al. Retzius-sparing robot-assisted radical prostatectomy vs the standard approach: a systematic review and analysis of comparative outcomes. BJU Int. 2020;125(1):8– 16. doi:10.1111/bju.14887

8. Viani G, Hamamura AC, Faustino AC. Intensity modulated radiotherapy (IMRT) or conformational radiotherapy (3D-CRT) with conventional fractionation for prostate cancer: Is there any clinical difference? Int Braz J Urol Off J Braz Soc Urol. 2019 Dec 17;45(6):1105–12. doi:10.1590/S1677-5538.IBJU.2018.0842 PubMed PMID: 31808397; PubMed Central PMCID: PMC6909869.

9. Tanaka N. The oncologic and safety outcomes of low-dose-rate brachytherapy for the treatment of prostate cancer. Prostate Int. 2023 Sep 1;11(3):127–33. doi:10.1016/j.prnil.2023.01.004

10. Chen RC, Basak R, Meyer AM, Kuo TM, Carpenter WR, Agans RP, et al. Association between choice of radical prostatectomy, external beam radiotherapy, brachytherapy, or active surveillance and patient-reported quality of life among men with localized prostate cancer. JAMA. 2017 Mar 21;317(11):1141–50. doi:10.1001/jama.2017.1652 PubMed PMID: 28324092; PubMed Central PMCID: PMC6284802.

11. Chien GW, Slezak JM, Harrison TN, Jung H, Gelfond JS, Zheng C, et al. Health-related quality of life outcomes from a contemporary prostate cancer registry in a large diverse population. BJU Int. 2017;120(4):520–9. doi:10.1111/bju.13843

12. van Stam MA, Aaronson NK, Bosch JLHR, Kieffer JM, van der Voort van Zyp JRN, Tillier CN, et al. Patient-reported Outcomes Following Treatment of Localised Prostate Cancer and Their Association with Regret About Treatment Choices. Eur Urol Oncol. 2020 Feb 1;3(1):21–31. doi:10.1016/j.euo.2018.12.004

13. Hoffman KE, Penson DF, Zhao Z, Huang LC, Conwill R, Laviana AA, et al. Patient-Reported Outcomes Through 5 Years for Active Surveillance, Surgery, Brachytherapy, or External Beam Radiation With or Without Androgen Deprivation Therapy for Localized Prostate Cancer. JAMA. 2020 Jan 14;323(2):149–63. doi:10.1001/jama.2019.20675 PubMed PMID: 31935027; PubMed Central PMCID: PMC6990712.

14. Barocas DA, Alvarez J, Resnick MJ, Koyama T, Hoffman KE, Tyson MD, et al. Association between radiation therapy, surgery, or observation for localized prostate cancer and patient-reported outcomes after 3 years. JAMA. 2017 Mar 21;317(11):1126–40. doi:10.1001/jama.2017.1704 PubMed PMID: 28324093; PubMed Central PMCID: PMC5782813.

15. Al Hussein Al Awamlh B, Wallis CJD, Penson DF, Huang LC, Zhao Z, Conwill R, et al. Functional Outcomes After Localized Prostate Cancer Treatment. JAMA. 2024 Jan 23;331(4):302–17. doi:10.1001/jama.2023.26491 PubMed PMID: 38261043; PubMed Central PMCID: PMC10807259.

16. Corsini C, Bergengren O, Carlsson S, Garmo H, Hjelm-Eriksson M, Fransson P, et al. Patient-reported Side Effects 1 Year After Radical Prostatectomy or Radiotherapy for Prostate Cancer: A Register-based Nationwide Study. Eur Urol Oncol. 2024 Jun;7(3):605–13. doi:10.1016/j.euo.2023.12.007 PubMed PMID: 38233329; PubMed Central PMCID: PMC11102330.

17. van Stam MA, Aaronson NK, Bosch JLHR, Kieffer JM, van der Voort van Zyp JRN, Tillier CN, et al. Patient-reported Outcomes Following Treatment of Localised Prostate Cancer and Their Association with Regret About Treatment Choices. Eur Urol Oncol. 2020 Feb 1;3(1):21–31. doi:10.1016/j.euo.2018.12.004

18. Hoffman KE, Penson DF, Zhao Z, Huang LC, Conwill R, Laviana AA, et al. Patient-Reported Outcomes Through 5 Years for Active Surveillance, Surgery, Brachytherapy, or External Beam Radiation With or Without Androgen Deprivation Therapy for Localized Prostate Cancer. JAMA. 2020 Jan 14;323(2):149–63. doi:10.1001/jama.2019.20675 PubMed PMID: 31935027; PubMed Central PMCID: PMC6990712.

19. Zamora V, Garin O, Suárez JF, Gutiérrez C, Guedea F, Cabrera P, et al. Comparative effectiveness of new treatment modalities for localized prostate cancer through patient-reported outcome measures. Clin Transl Radiat Oncol. 2023 Oct 29;44:100694. doi:10.1016/j.ctro.2023.100694 PubMed PMID: 38021091; PubMed Central PMCID: PMC10663757.

20. Ferrer M, Garin O, Pera J, Prats JM, Mendivil J, Alonso J, et al. Evaluación de la calidad de vida de los pacientes con cáncer de próstata localizado: validación de la versión española del cuestionario EPIC. Med Clínica. 2009 Feb 7;132(4):128–35. doi:10.1016/j.medcli.2008.01.001

21. Szymanski KM, Wei JT, Dunn RL, Sanda MG. Development and Validation of an Abbreviated Version of the Expanded Prostate Cancer Index Composite Instrument (EPIC-26) for Measuring Health-Related Quality of Life Among Prostate Cancer Survivors. Urology. 2010 Nov;76(5):1245–50. doi:10.1016/j.urology.2010.01.027 PubMed PMID: 20350762; PubMed Central PMCID: PMC3152317.

22. Alonso J, Prieto L, Antó JM. [The Spanish version of the SF-36 Health Survey (the SF-36 health questionnaire): an instrument for measuring clinical results]. Med Clin (Barc). 1995 May 27;104(20):771–6. PubMed PMID: 7783470.

23. Vilagut G, Ferrer M, Rajmil L, Rebollo P, Permanyer-Miralda G, Quintana JM, et al. [The Spanish version of the Short Form 36 Health Survey: a decade of experience and new developments]. Gac Sanit. 2005;19(2):135–50. doi:10.1157/13074369 PubMed PMID: 15860162.

24. ResearchGate [Internet]. [cited 2025 Aug 6]. How to Score Version 2 of the SF-36® Health Survey | Request PDF. Available from: https://www.researchgate.net/publication/246394689_How_to_Score_Version_2_of_the_SF-36R_Health_Survey

25. Wei JT, Dunn RL, Litwin MS, Sandler HM, Sanda MG. Development and validation of the expanded prostate cancer index composite (EPIC) for comprehensive assessment of health-related quality of life in men with prostate cancer. Urology. 2000 Dec 1;56(6):899–905. doi:10.1016/S0090-4295(00)00858-X

26. Skolarus TA, Dunn RL, Sanda MG, Chang P, Greenfield TK, Litwin MS, et al. Minimally important difference for the Expanded Prostate Cancer Index Composite Short Form. Urology. 2015 Jan;85(1):101–5. doi:10.1016/j.urology.2014.08.044 PubMed PMID: 25530370; PubMed Central PMCID: PMC4274392.

27. Li F, Li F. Propensity Score Weighting for Causal Inference with Multiple Treatments [Internet]. arXiv; 2019 [cited 2025 Oct 19]. Available from: http://arxiv.org/abs/1808.05339 doi:10.48550/arXiv.1808.05339

28. Dalela D, Jeong W, Prasad MA, Sood A, Abdollah F, Diaz M, et al. A Pragmatic Randomized Controlled Trial Examining the Impact of the Retzius-sparing Approach on Early Urinary Continence Recovery After Robot-assisted Radical Prostatectomy. Eur Urol. 2017 Nov 1;72(5):677–85. doi:10.1016/j.eururo.2017.04.029

29. Qiu X, Li Y, Chen M, Xu L, Guo S, Marra G, et al. Retzius-sparing robot-assisted radical prostatectomy improves early recovery of urinary continence: a randomized, controlled, single-blind trial with a 1-year follow-up. BJU Int. 2020;126(5):633–40. doi:10.1111/bju.15195

30. Luckenbaugh AN, Wallis CJD, Huang LC, Wittmann D, Klaassen Z, Zhao Z, et al. Association between Treatment for Localized Prostate Cancer and Mental Health Outcomes. J Urol. 2022 May. Located at: Philadelphia, PA. doi:10.1097/JU.0000000000002370

31. Brunckhorst O, Liszka J, James C, Fanshawe JB, Hammadeh M, Thomas R, et al. Mental well-being in prostate cancer: A multi-institutional prospective cohort study. BJUI Compass. 2025;6(6):e70040. doi:10.1002/bco2.70040

32. Martin NE, Massey L, Stowell C, Bangma C, Briganti A, Bill-Axelson A, et al. Defining a Standard Set of Patient-centered Outcomes for Men with Localized Prostate Cancer. Eur Urol. 2015 Mar 1;67(3):460–7. doi:10.1016/j.eururo.2014.08.075

